# Near-infrared spectroscopy reveals asymmetric hemispheric blood supply during aortic arch surgery

**DOI:** 10.64898/2026.11.09.26367255

**Authors:** Yasin El Mokhtari, F. Konrad Schumacher, Justyna Podlasek, Florian Meissner, Alina Hiller, Domagoj Damjanovic, Frank Humburger, Fabio Brizzi, Helena Kirr, Tim Berger, Maximilian Kreibich, Sven Maier, Marius Schimmel, Roman Gottardi, Horst Urbach, Josef M. Unterrainer, Martin Czerny, Stoyan Kondov, Christoph P. Kaller

**Author notes:** Yasin El Mokhtari and F. Konrad Schumacher contributed equally to this work. **Corresponding authors** — Yasin El Mokhtari, Department of Cardiac, Thoracic, Vascular Surgery and Aortic Medicine, University Heart Center, Südring 15, 79189 Bad Krozingen, Germany; Christoph P. Kaller, Neurocenter, Breisacher Str. 64, 79106 Freiburg, Germany.

## Abstract

**Objectives:** Bilateral and trilateral selective antegrade cerebral perfusion (SACP) aim to ensure symmetric hemispheric perfusion during aortic arch surgery, which current monitoring cannot directly assess. As a step towards real-time monitoring, we assessed the feasibility of multichannel contrast-enhanced near-infrared spectroscopy (CE-NIRS) using indocyanine green and whether hemispheric bolus timing matched expectations for each perfusion configuration.

**Methods:** In this prospective, offline-analysed pilot study, 10 patients undergoing aortic arch surgery (9 frozen elephant trunk) received 94 boluses across 10 perfusion configurations. The interhemispheric difference in time to peak (ΔTTP) was compared with the pre-bypass baseline using a linear mixed model with Holm correction, alongside cerebral oximetry; the relationship to postoperative cerebral injury was explored post hoc.

**Results:** 92 of 94 boluses yielded a bilateral TTP. Contrary to expectation, the left hemisphere peaked later under bilateral (+7.2 s, 95% CI 3.4–11.0; p = 0.003) and trilateral SACP (+8.7 s, 2.5–14.8; p = 0.031). With both left-sided arch vessels on body perfusion, the asymmetry reversed (−7.0 s, −11.7 to −2.3; p = 0.025). ΔTTP detected 6 of 9 configuration effects, whereas oximetry detected none after correction. The three patients with cerebral injury showed the three most pronounced reversals (exact p = 0.100) but were also those managed trilaterally.

**Conclusions:** Multichannel CE-NIRS is feasible during aortic arch surgery, without indicator-related adverse events. Bilateral and trilateral SACP did not secure symmetric indicator delivery; the configuration-dependent asymmetry and its reversal point to two steals within the perfusion circuit that oximetry missed. Their clinical relevance requires prospective confirmation.

## Introduction

Total arch replacement with the frozen elephant trunk (FET) technique has become a widely adopted strategy for complex arch and proximal descending aortic disease.^1^ During the operation, the brain is supplied in a sequence of configurations: by the native circulation, the cardiopulmonary bypass (CPB) circuit, selective antegrade cerebral perfusion (SACP) through cannulae in one, two, or three arch vessels (unilateral, bilateral, or trilateral SACP), and again by body perfusion as the anastomoses are completed. Stroke remains a decisive complication of FET repair^2^: in 250 repairs at our centre (2013–2020), 16.8% of patients had a symptomatic stroke, mostly embolic.^3^

Which configuration protects the brain best remains unresolved. Meta-analyses comparing unilateral and bilateral SACP found no significant differences in mortality and neurological outcome,^4^ less neurological injury^5^ versus more transient deficits^6^ with unilateral perfusion. Furthermore, in a propensity-matched series, trilateral and bilateral SACP did not differ in FET repair.^7^ Recommendations for bilateral or trilateral perfusion rest on anatomical variants of the circle of Willis and on the anticipated duration of SACP.^1^ Under bilateral SACP, delivered flow was linked to saturation and outcome,^8^ but whether flow reaches both hemispheres equally is not measured directly: oximetry, transcranial Doppler, and processed electroencephalography each capture only part of cerebral physiology,^9^ with bilateral oximetry remaining the recommended standard.^1^

Multichannel contrast-enhanced near-infrared spectroscopy (CE-NIRS) with indocyanine green (ICG) bolus tracking can provide this intraoperative measurement for each hemisphere: the time to peak (TTP) of the bolus has previously been used to detect perfusion deficits in acute stroke via interhemispheric differences.^10^ In steno-occlusive cerebrovascular disease, multichannel maps have demonstrated delays over the affected hemisphere,^11^ and the side ratios of bolus kinetics were found to be consistent with positron emission tomography.^12^ CE-NIRS has been applied in humans during CPB,^13^ but only in piglets during SACP.^14^ Multichannel CE-NIRS samples multiple sites across both hemispheres rather than a single frontal field, as multichannel oximetry already has during aortic arch replacement.^15^ Because each bolus is resolved within its first passage, the measurement could in the future be displayed intraoperatively in real time, while the perfusion configuration can still be adjusted.

Before such real-time monitoring can be used intraoperatively, its feasibility and potential added value must be demonstrated. In this offline-analysed pilot study, we therefore asked: first, whether multichannel CE-NIRS is feasible throughout FET repair; second, whether the interhemispheric TTP difference of each configuration matches the symmetry it is intended to provide (Table S2); and third, how this measurement relates to standard cerebral oximetry and postoperative cerebral injury.

## Methods

### Study design and patients

This prospective single-centre observational pilot study was conducted at the Department of Cardiac, Thoracic, Vascular Surgery and Aortic Medicine, Medical Center – University of Freiburg, Germany, between September 2022 and October 2024. It was approved by the institutional ethics committee (vote 335/20), and conducted in accordance with the Declaration of Helsinki; all patients provided written informed consent. Ten adults undergoing aortic arch surgery with SACP were enrolled when the measurement team and equipment were available: nine for FET repair and one for hemiarch replacement (Table 1; full characteristics in Table S1). As a pilot study, no formal sample size calculation was performed. Exclusion criteria were known allergy to ICG, severe pre-existing neurological impairment precluding postoperative assessment, and emergency surgery. The measurement did not alter cerebral protection or surgical technique.

**Table 1.** Patient and operative characteristics (*n* = 10). Data are *n* (%) or mean ± SD. CPB: cardiopulmonary bypass; FET: frozen elephant trunk; LVEF: left ventricular ejection fraction.

| Characteristic | Value |
|---|---|
| **Demographics** |  |
| Age at surgery, years | 66.6 ± 10.7 |
| Female sex | 3 (30%) |
| Body mass index, kg/m² | 27.9 ± 7.0 |
| **Comorbidities** |  |
| Hypertension | 10 (100%) |
| Diabetes mellitus | 2 (20%) |
| Coronary artery disease | 1 (10%) |
| LVEF <50% | 2 (20%) |
| Preoperative atrial fibrillation | 2 (20%) |
| Previous stroke | 1 (10%) |
| Supra-aortic vessel stenosis >65% | 4 (40%) |
| Supra-aortic vessel dissection | 3 (30%) |
| **Aortic pathology** |  |
| Native arch aneurysm | 4 (40%) |
| Post-dissection aneurysm | 1 (10%) |
| Postoperative aneurysm progression | 5 (50%) |
| Previous aortic surgery | 6 (60%) |
| **Operation** |  |
| Isolated FET | 3 (30%) |
| Additional cardiac procedure | 5 (50%) |
| Redo cardiac surgery | 7 (70%) |
| Urgent operation | 2 (20%) |
| Cerebral perfusion: bilateral / trilateral | 6 (60%) / 4 (40%) |
| EuroSCORE II, % | 5.7 ± 4.8 |
| Operation time, min | 404.2 ± 80.7 |
| CPB time, min | 237.5 ± 47.7 |
| Cross-clamp time, min | 118.9 ± 30.2 |
| Lowest body temperature, °C | 24.4 ± 2.1 |

### Multichannel CE-NIRS and indicator protocol

Two cascaded continuous-wave NIRSport2 devices (NIRx Medizintechnik, Berlin, Germany; 760 and 850 nm, 5.1 Hz) provided 32 sources and 32 detectors, distributed over a 5 × 5 array on the frontotemporal region of each hemisphere (centred approximately between F7–P7 and F8–P8; Fig. 1), two shoulder probes as extracerebral references, and 8-mm short-separation channels. The head arrays yielded source–detector separations of 18, 40 and 54 mm; the analysis used the 40-and 54-mm channels, which are sensitive to cortical tissue.^16^ ICG (Diagnostic Green, 0.1 mg/kg, 1.67 mg/mL) was injected over 2 s by a power injector (MEDRAD Mark 7 Arterion, Bayer) under software control that time-stamped each injection in the recording. Boluses were administered through the central venous catheter before and after CPB, and into the CPB circuit otherwise. Regional cerebral oxygen saturation (rSO_2_) was recorded bilaterally with a clinical oximeter (INVOS 5100C; Medtronic, Minneapolis, MN, USA). Hardware, montage, and preprocessing are detailed in the Supplementary Methods.

**Figure 1.**
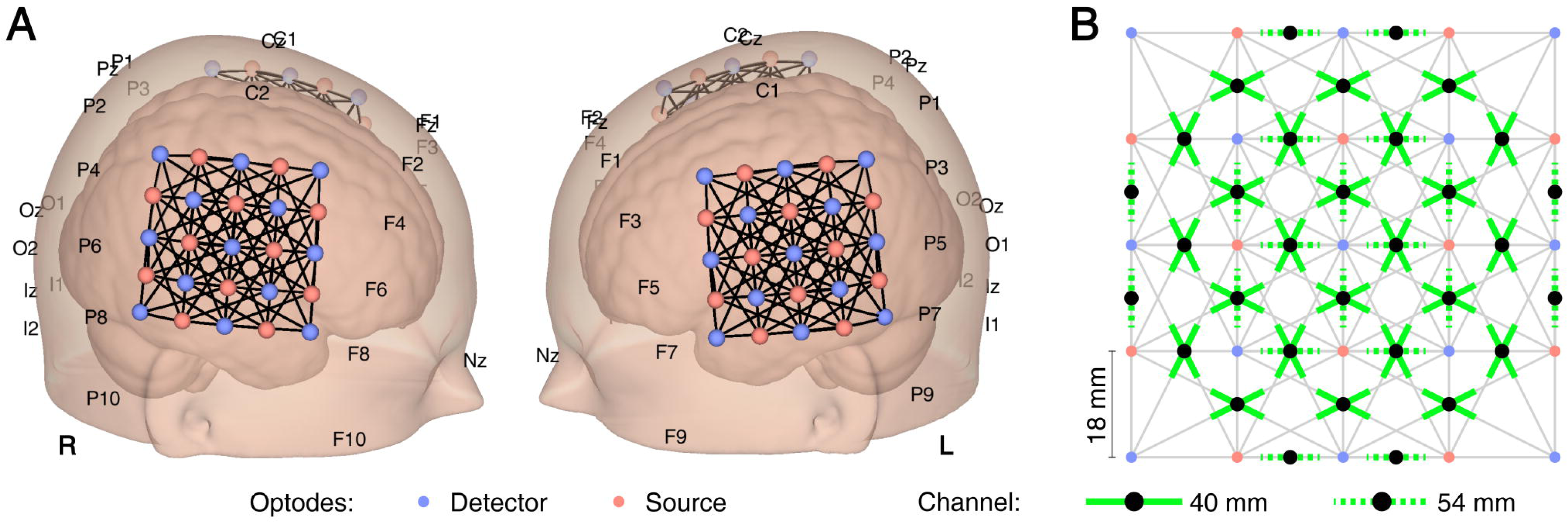
CE-NIRS montage (schematic illustration). **(A)** Sources (red) and detectors (blue) of the two head arrays, with landmarks of the 10–20 system. **(B)** Analysed 40- and 54-mm channels of one hemisphere (*n* = 48 and 20), drawn as segments along their source–detector axis. CE-NIRS: contrast-enhanced near-infrared spectroscopy. *Alt text:* Two three-dimensional renderings of a head, viewed from the right and from the left, each with a square array of near-infrared light sources (red) and detectors (blue) over the lateral aspect of the hemisphere; lines between them mark the measurement channels, and positions of the 10–20 system are labelled. A second panel shows the grid of one array, with neighbouring optodes 18 mm apart and the analysed channels at 40 mm (solid green) and 54 mm (dashed green) source–detector distance.

### Surgical technique and perfusion configurations

CPB was established via the right subclavian artery (RSA), which also provided the primary SACP inflow. Bilateral SACP added direct cannulation of the left common carotid artery (LCCA); trilateral SACP additionally perfused the left subclavian artery (LSA) or the left vertebral artery. Six patients were managed bilaterally and four trilaterally. The selective lines and the body-perfusion line branched from the arterial line through T- and Y-connectors (Fig. 2). SACP was delivered by a separate roller pump during simultaneous lower body perfusion.^17^

**Figure 2.**
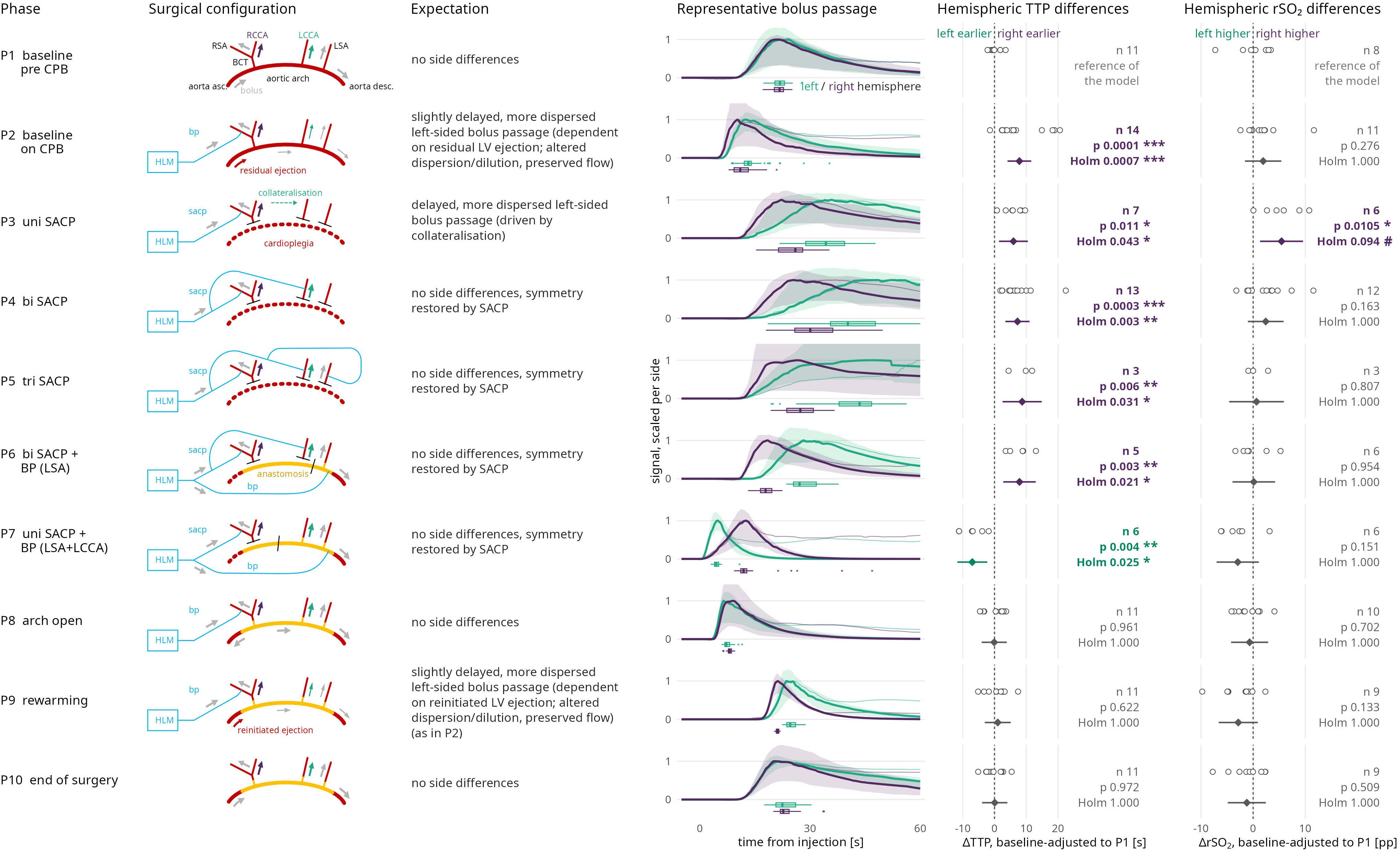
Interhemispheric differences in TTP and rSO_2_ across the ten perfusion configurations. Rows: configurations P1–P10 in surgical order. Columns show the circuit, the specified expectation (Table S2), a representative bolus (per hemisphere, median of the fitted curves with interquartile band; thin line, measured signal; box plot, channel peak times), and the side differences in TTP and rSO_2_: each bolus as an open circle, the contrast against P1 as a diamond with 95% CI, with *n*, *p* and Holm-adjusted *p*. Values are adjusted for patient offset and P1 level (linear mixed model; Supplementary Methods). On both axes a negative value marks the left hemisphere as ahead (earlier peak, higher saturation). BCT: brachiocephalic trunk; BP: body perfusion; CPB: cardiopulmonary bypass; HLM: heart–lung machine; LCCA/RCCA: left/right common carotid artery; LSA/RSA: left/right subclavian artery; rSO_2_: regional oxygen saturation; SACP: selective antegrade cerebral perfusion; TTP: time to peak. *Alt text:* Figure with one row per perfusion configuration in surgical order (P1 to P10). Each row shows a schematic of the aortic arch and perfusion circuit, the expected side difference, representative left- and right-hemispheric bolus curves, and dot plots of the interhemispheric differences in time to peak and in regional oxygen saturation with model estimates and P values. Time to peak was later on the left from P2 to P6, including bilateral and trilateral selective perfusion, and reversed to left-earlier when both left-sided arch vessels were on body perfusion (P7); saturation differences were small and none remained significant after correction.

Each bolus was assigned to one of ten perfusion configurations (Fig. 2, Table S2): baseline before (P1) and on CPB (P2); unilateral (P3), bilateral (P4), and trilateral SACP (P5); bilateral SACP with the LSA on body perfusion (P6); unilateral SACP with the LSA and LCCA on body perfusion (P7); arch open (P8); rewarming (P9); and end of surgery (P10). For each configuration, the expected interhemispheric TTP difference was specified based on the circuit and the intended function of the configuration, in particular, symmetry under bilateral and trilateral SACP.

### Signal analysis

After adaptive filtering that preserves the timing and height of the bolus peak (Supplementary Methods), the ICG time course of each channel was fitted with a biphasic growth model that separates the first pass from recirculation. TTP is the time from injection to the maximum of the fitted curve; the rise time is the interval from its onset to that maximum. TTP thus describes how early and how dispersed the indicator reaches the tissue, not tissue perfusion itself. A channel entered the analysis if it met two acceptance criteria: R² ≥ 0.9 and a rise time below 59 s, excluding channels whose maximum occurs during recirculation. The hemispheric TTP of a bolus is the median over its accepted channels. The primary endpoint is the interhemispheric difference ΔTTP = TTP(left) − TTP(right). The rSO_2_ difference was calculated as right − left, so that a positive value of either quantity identifies the left hemisphere as disadvantaged (later peak, lower saturation). Channel yield, the proportion of montaged channels passing both acceptance criteria, is defined in the Supplementary Methods.

### Outcome

Following clinical standards, cerebral computed tomography was performed when a patient exhibited new neurological symptoms after surgery until discharge. Postoperative cerebral injury was defined as a new ischaemic or hypoxic lesion on this imaging.

### Statistical analysis

ΔTTP was analysed using a linear mixed model with perfusion configuration as a fixed effect and a random intercept per patient, fitted by restricted maximum likelihood. Each configuration was contrasted against P1 with 95% confidence intervals (CI) and Holm correction across the nine contrasts. The patient intercept accounts for each patient’s native side asymmetry; because P1 serves as the reference level rather than being subtracted, the patient without a P1 bolus remained in the analysis. The rSO_2_ difference was analysed with the identical model. The association between rSO_2_ and TTP was estimated from bilateral SACP to arch completion (P4– P8) using a linear mixed model of hemisphere-specific rSO_2_ on TTP with a TTP × hemisphere interaction, configuration as a fixed effect, and random intercepts for patient and bolus, so that the slope reflects variation within a configuration. In P7, patients with and without cerebral injury were compared using an exact Wilcoxon rank-sum test on ΔTTP; with three patients per group, the smallest attainable two-sided *p*-value is 0.100. Holm correction was applied within each family of tests but not between families (Supplementary Methods); the association and the outcome comparison are exploratory and hypothesis-generating. Analyses used R 4.5.3^18^ with lme4, lmerTest, and emmeans (Supplementary Methods).

## Results

### Feasibility

All ten patients completed the protocol. Ninety-four ICG boluses were administered without an adverse event attributable to the indicator, and 92 (97.9%) yielded a valid TTP in both hemispheres. The two exceptions occurred in one patient, with loss of the left-hemispheric signal in P7 and both hemispheres falling below the acceptance criteria in P6 (Supplementary Results). Coverage of the configurations was uneven by design and surgical course: P2, P4, P8, and P10 were captured in all ten patients, trilateral SACP (P5) in three of the four patients managed trilaterally (Table S3). Overall, 69.0% of the montaged 40- and 54-mm channels met both acceptance criteria.

### ΔTTP across perfusion configurations

Before CPB (P1), the bolus peaked almost simultaneously in both hemispheres (raw median ΔTTP +1.7 s, IQR +0.8 to +2.6 s). On CPB before SACP (P2), ΔTTP rose by 7.8 s (95% CI 4.1 to 11.6; Holm-adjusted *p* = 0.001), and once each patient’s P1 asymmetry was removed, the left hemisphere peaked first in only 1 of 14 boluses (none unadjusted). The right-sided lead persisted under unilateral SACP (P3; +6.0 s, 1.4 to 10.5; *p* = 0.043) and, contrary to the purpose of escalating cannulation, under bilateral (P4; +7.2 s, 3.4 to 11.0; *p* = 0.003) and trilateral SACP (P5; +8.7 s, 2.5 to 14.8; *p* = 0.031) and after transfer of the LSA to body perfusion (P6; +7.9 s, 2.8 to 13.0; *p* = 0.021; Fig. 2). Once each patient’s native side asymmetry in P1 was removed, all 21 boluses from P4 to P6 showed a right-sided lead (20 of 21 unadjusted).

When both left-sided arch vessels were on body perfusion and the RSA remained the only SACP line (P7), the asymmetry reversed (−7.0 s, −11.7 to −2.3; *p* = 0.025). The left hemisphere peaked first in five of six boluses; relative to each patient’s baseline, this reversal was present in all six patients (Fig. 3A; an individual course in Fig. 3D). After completion of the arch, ΔTTP returned to baseline (P8–P10: −0.1, +1.0 and +0.1 s; Holm-adjusted *p* = 1.000).

**Figure 3.**
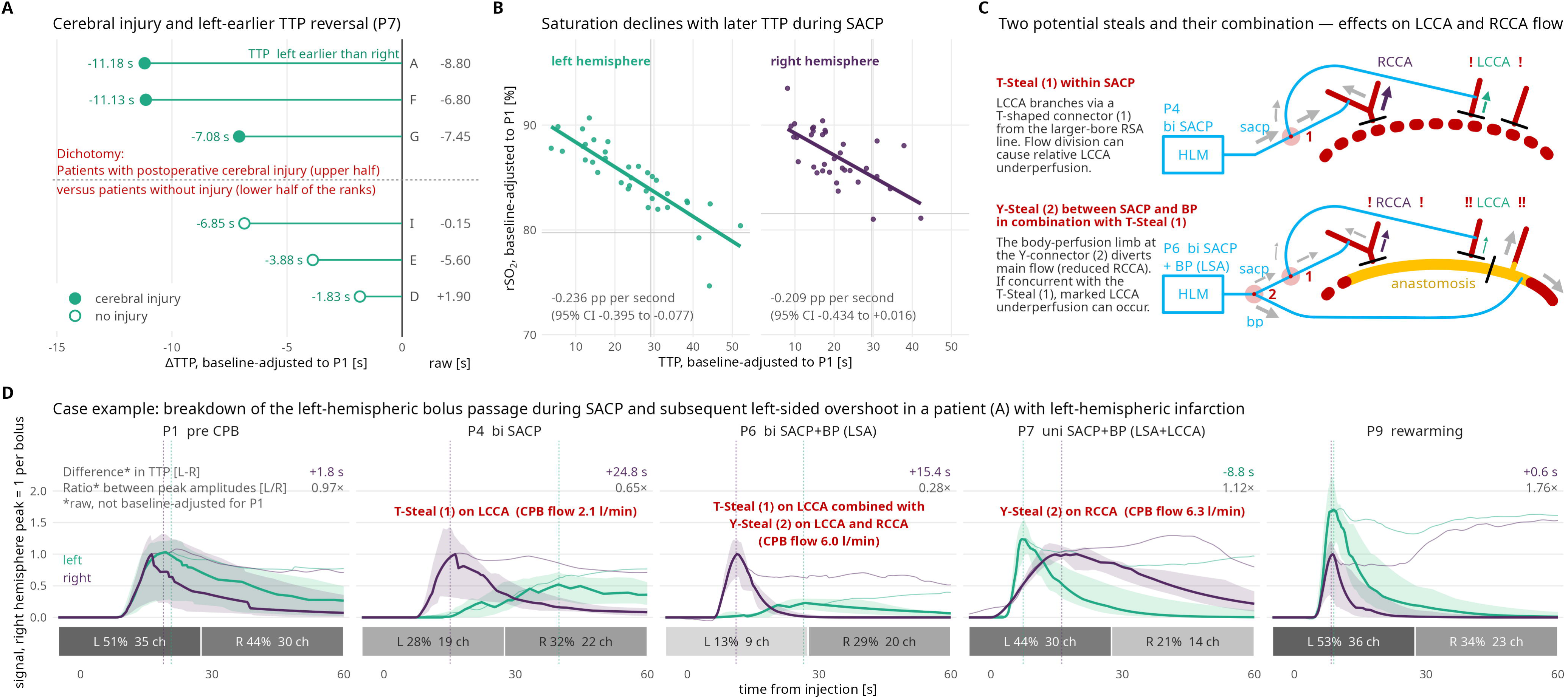
TTP reversal with both left-sided arch vessels on body perfusion (P7), its relation to rSO_2_, and the proposed mechanisms. **(A)** ΔTTP in P7, one bolus per patient (*n* = 6), adjusted for patient offset and P1 level; raw values beside the panel. Filled symbols: postoperative cerebral injury. **(B)** rSO_2_ against TTP during SACP (P4–P7) and after arch completion (P8), one point per bolus and hemisphere, adjusted for patient, bolus and configuration. **(C)** Proposed T-steal (1) at the T-connector of the LCCA line and Y-steal (2) at the Y-connector between selective and body perfusion. **(D)** Bolus series of patient A: median of the fitted curves with interquartile band; thin lines, measured signal; dashed lines, hemispheric TTP; numbers give ΔTTP and the left-to-right peak ratio, tiles the channel yield per side. Curves are scaled to the right-hemispheric peak of each bolus, so heights compare only as ratios. pp: percentage points; other abbreviations as in Figure 2. *Alt text:* Four panels. (A) Ranked interhemispheric time-to-peak differences in configuration P7 for six patients; the three patients with postoperative cerebral injury (filled symbols) show the largest left-earlier reversals. (B) Scatter plots for the left and right hemisphere in which regional oxygen saturation declines with later time to peak, with regression lines. (C) Schematics of the perfusion circuit illustrating two proposed steals, at the T-connector of the left common carotid line and at the Y-connector between selective and body perfusion. (D) Bolus curves of one patient with left-hemispheric infarction across five configurations: the left-hemispheric bolus is delayed and flattened during selective perfusion, peaks earlier than the right in P7 and higher than the right during rewarming.

### Cerebral oximetry

The interhemispheric rSO_2_ difference (right − left), analysed with the same model, differed from baseline only nominally under unilateral SACP (P3; +5.4 percentage points, 1.3 to 9.6; unadjusted *p* = 0.010), and no contrast survived Holm correction (smallest adjusted *p* = 0.094; Supplementary Results). Within the configurations in which the arch vessels change between selective and body perfusion (P4–P8), excluding the circulatory states before SACP and during rewarming, rSO_2_ nevertheless fell with increasing TTP: by −0.24 percentage points per second on the left (95% CI −0.40 to −0.08; *p* = 0.004) and by −0.21 on the right, where it is reported as a trend (−0.43 to +0.02; *p* = 0.068), without a difference between hemispheres (interaction *p* = 0.71; Fig. 3B). Readings at the device ceiling of 95% (11.4% in this window) bias these slopes towards zero.

### Channel yield

Channel yield fell on both sides when selective and body perfusion ran in parallel (P6: 33% left, 54% right; P7: 43% and 66%; P4: 57% and 83%), and its side difference followed the ΔTTP pattern (ρ = 0.70; Supplementary Results, Supplementary Fig. S1).

### Postoperative cerebral injury

Three patients had postoperative cerebral injury on computed tomography: a left-hemispheric infarction, a right-hemispheric infarction, and a malignant right middle cerebral artery infarction with extensive, presumably hypoxic, left-hemispheric oedema (Table S5). All three had a bilateral value in P7 and showed the three most pronounced ΔTTP reversals. In contrast, the three patients without cerebral injury ranked fourth to sixth on both the adjusted and raw scales (exact Wilcoxon *p* = 0.100, the smallest value attainable with three patients per group; Fig. 3A). However, outcome and cannulation strategy coincided exactly in this configuration: the three patients with cerebral injury were those managed trilaterally, so the separation cannot be attributed to outcome independently of strategy. Because the configuration and comparison were chosen post hoc, this finding is hypothesis-generating.

## Discussion

### Principal findings

This pilot study shows that multichannel CE-NIRS is feasible throughout aortic arch surgery with SACP, with a bilateral TTP in 97.9% of boluses and no indicator-related adverse events. Its principal finding is that neither bilateral nor trilateral SACP restored the interhemispheric symmetry of indicator delivery that these strategies are meant to secure: the left hemisphere peaked later in 20 of 21 boluses (all 21 when adjusted to P1), by 7 to 9 s relative to this baseline. The asymmetry depended on the circuit configuration rather than on the cannulation strategy alone — it reversed when both left-sided arch vessels were on body perfusion and disappeared once the arch was completed; the left-sided delay expected during rewarming was absent, probably because left-ventricular ejection had not yet resumed (Supplementary Discussion).

Such a persistent asymmetry under escalated cannulation may help explain why meta-analyses have not shown consistent outcome differences between unilateral and bilateral SACP.^4–6^

### Flow competition at the circuit junctions

We propose that both patterns arise from flow competition at the junctions of the circuit, where flow follows the path of least resistance (Fig. 3C). Under bilateral and trilateral SACP, the left common carotid cannula branches through a T-connector from the larger-bore line to the right subclavian artery; the straight passage favours the right side (T-steal). This is consistent with mock-loop and computational data in which left-sided inflow fell by about half across cannulae of different resistance,^19^ and with an early multichannel oxygenation recording in which the hemispheric balance differed between two patients with different LCCA cannula sizes.^15^ Once both left-sided arch vessels were transferred to body perfusion (P7), the right hemisphere depended on the SACP limb alone, while the left was supplied through the body-perfusion limb, which diverges from the arterial line at a Y-connector and offers lower resistance; the disadvantage then shifted to the right (Y-steal). The SACP roller pump, not run fully occlusively, does not fix the flow in its limb (Supplementary Discussion). Between these two states, in P6, the LSA was already on body perfusion while the LCCA was still fed through the T-connector, so that both steals could act on the left hemisphere. The strength of either steal varied widely between patients within the same configuration, consistent with the configuration creating the condition for a steal and patient factors such as collateral capacity determining its effect (Supplementary Discussion).

A longer line to the left hemisphere would also delay its peak, but by our estimate at the T-connector only by about 1 s (Supplementary Discussion), and it would shift the curve without reducing the signal or the saturation. Instead, the asymmetry was accompanied by a loss of left-sided channel yield that followed ΔTTP across configurations, and a later peak was associated with a lower rSO_2_ within configurations. In our setting, flow in the individual limbs could not be measured because the connectors lay in the sterile field without flow sensors. Only the arterial pump flow, calculated from pump speed, was documented. The mechanism therefore remains an inference from the circuit topology and the kinetic bolus data.

### Comparison with cerebral oximetry

Standard oximetry did not detect the configuration effects: only one of nine contrasts reached nominal significance and none survived correction, compared with six of nine for ΔTTP. Saturation was not unrelated to indicator delivery, but a later TTP was associated with a lower rSO_2_ by only about 0.2 percentage points per second, so that delays of 7 to 9 s correspond to about 2 percentage points. Single-region oximetry samples a small frontal field, is sensitive to extracerebral signal^9^ and has missed focal ischaemia confirmed by other modalities.^20^ In an exploratory analysis of a randomised trial, desaturation was unrelated to new lesions on diffusion-weighted imaging,^21^ and in 166 FET repairs rSO_2_ was of limited value for predicting stroke,^22^ although relative desaturation identified stroke in an earlier series.^23^ TTP and rSO_2_ thus capture different aspects of supply, and the oximetric monitor can miss a delivery asymmetry during SACP.

### Cerebral injury as a hypothesis

The three patients with postoperative cerebral injury showed the three strongest P7 reversals. This observation is post hoc, rests on three-against-three patients, and is confounded: in this configuration, the patients with cerebral injury were exactly those managed trilaterally, a strategy chosen because of the posterior circulation or variants of the circle of Willis, which are common.^24^ The magnitude of the reversal may therefore index the strength of the Y-steal, limited collateral compensation, or both. How closely the lesion matched the recorded field differed among the three patients, and in one case, a mechanical cause at the graft anastomosis must be considered (Table S5; Supplementary Discussion).

## Limitations

The cohort is small, coverage of the configurations was uneven, and TTP reflects the kinetics of delivery, not flow. Postoperative imaging was obtained only in symptomatic patients, so clinically silent lesions cannot be excluded, and whether the injuries arose intraoperatively or after surgery is unknown. The concurrent clinical oximetry reduced left-sided channel yield, and the association with cerebral injury was examined after inspection of the data.

## Conclusions

Multichannel CE-NIRS reveals a configuration-dependent asymmetry of cerebral delivery during SACP that escalated cannulation does not remove and that standard oximetry does not detect. A prospective multicentre study should define the configurations of interest and the reversal metric in advance, include systematic postoperative diffusion-weighted imaging, and combine the measurement with flow measurements in the circuit limbs^25^ or mock-loop studies of the connectors. A future real-time display would allow the OR team to recognise an emerging steal while the perfusion configuration can still be adjusted.

## Supporting information

STROBE Checklist

## Acknowledgements

During the preparation of this work, the authors used Claude Opus 5.5 (Anthropic PBC, San Francisco, CA, USA) to write code for data analyses and figure creation and for language editing of the manuscript. Following the use of this tool, the authors thoroughly reviewed and edited the content as necessary and take full responsibility for the content of the published article, including the statistical analysis of the data.

## Funding

This work was supported by intramural funding from the Faculty of Medicine, University of Freiburg (award number 2023/A6-Kal). The funder had no role in study design, analysis or reporting.

## Conflict of interest

M.C. and R.G. are shareholders of Ascense Medical and TEVAR, outside the submitted work. S.K. is a consultant for Artivion, SB-Kawasumi Laboratories, Terumo and Lifetech. T.B. received speaking honoraria from Artivion. M.K. received speaking honoraria from Terumo Aortic and Artivion. H.U. received honoraria for lectures from Bayer, Biogen, GE, Eisai, Mbits and Lilly and is co-editor of Clinical Neuroradiology. F.K.S. and C.P.K. are named inventors on invention disclosures in the field of NIRS that are unrelated to the method reported here. The other authors declare no conflicts of interest.

## Author contributions

Conceptualization: FKS, MC, SK, CPK. Methodology: YEM, FKS, DD, FH, SM, RG, MC, SK, CPK. Software: FKS, CPK. Validation: YEM, FKS, AH, MS, CPK. Formal analysis: FKS, CPK. Investigation: YEM, FKS, JP, AH, DD, FH, FB, TB, MK, SM, MS, RG, MC, SK, CPK. Resources: HU, JMU, MC, CPK. Data curation: YEM, FKS, FM, AH, FB, CPK. Writing – original draft: YEM, FKS, CPK. Writing – review & editing: YEM, FKS, FM, SM, HU, JMU, MC, SK, CPK. Visualization: FKS, CPK. Supervision: HU, JMU, MC, SK, CPK. Project administration: YEM, FB, HK, JMU, MC, SK, CPK. Funding acquisition: HU, JMU, MC, CPK.

## Meeting presentation

Presented at the EACTS Annual Meeting 2026, Barcelona, Spain, 9 October 2026, as “Intraoperative Monitoring of Cerebral Perfusion During Selective Cerebral Perfusion in Frozen Elephant Trunk Repair Using Multichannel Near-Infrared Spectroscopy: Association with Postoperative Stroke”.

## Trial registration

Freiburg Clinical Studies Register (FRKS), FRKS005946.

## Data availability

The data underlying this article will be shared on reasonable request to the corresponding authors, subject to approval by the institutional ethics committee.

## Notes

### Author Declarations

The Ethics Committee of the University of Freiburg, Germany, gave ethical approval for this work (approval no. 335/20).

