## Supplementary Materials for "Near-infrared spectroscopy reveals asymmetric hemispheric blood supply during aortic arch surgery"

#### Supplementary methods

##### Study conduct

Inclusion criteria were aortic arch surgery with planned bilateral or trilateral selective antegrade cerebral perfusion, age ≥ 18 years and the capacity to give informed consent. The measurement was added to the institutional protocol without changing cannulation, perfusion or temperature management.

##### Contrast-enhanced near-infrared spectroscopy (CE-NIRS): hardware and montage

Two cascaded NIRSport2 devices (NIRx Medizintechnik, Berlin, Germany; Aurora recording software 2022.10.0-14) provided 32 LED sources (760 and 850 nm) and 32 avalanche photodiode detectors, sampled at 5.1 Hz. Each hemisphere carried a 5 × 5 checkerboard of 12 sources and 13 detectors with 18 mm spacing, yielding source–detector separations of 18, 40 and 54 mm; the inferior margins of the left and right arrays were centred approximately between the 10–20 positions (Jasper 1958) F7 and P7 and between F8 and P8, respectively, in standardised caps chosen by head circumference from sizes of 54 to 62 cm (EasyCap, Wörthsee, Germany). For extracerebral reference, 3D-printed probes with 4 sources each were attached to the shoulders. Short-channel bundles, each consisting of 16 fibres with two fibres per source optode, were read by dedicated detectors to provide 8-mm channels. These channels are predominantly sensitive to extracerebral haemodynamics (Strangman 2013). In two patients, two bundles served 8 sources per hemisphere and each shoulder probe had 2 detectors; in the other eight, three bundles served all 12 head sources per hemisphere, at the cost of one detector at the right shoulder. The head arrays and their 40- and 54-mm channels, on which all analyses rest, were identical in every recording; the shoulder probes and the short-separation channels were recorded but not analysed in the present study. The cap was fitted after the induction of anaesthesia and before draping, secured with a chin strap. The occiput was bedded on a gel headrest to avoid pressure injury and contact artefacts, and the optodes were shielded from ambient light with aluminium foil.

##### Signal conversion and indicator protocol

Changes in light intensity were converted to concentration changes of ICG and oxygenated haemoglobin using the modified Beer–Lambert law (Delpy 1988) in in-house R code. Because two wavelengths resolve only two chromophores, deoxygenated haemoglobin was not analysed, and boluses were administered only during haemodynamic stability. ICG (Diagnostic Green, Aschaffenburg, Germany) was dissolved in sterile water to 1.67 mg/mL; the volume for 0.1 mg/kg was rounded to the nearest millilitre and delivered at a constant rate over 2 s by a MEDRAD Mark 7 Arterion injector (Bayer Healthcare), triggered through its Imaging System Interface by custom MATLAB software (2023a; The MathWorks, Natick, MA, USA), which sent a synchronisation marker to the CE-NIRS recording via the Lab Streaming Layer (MATLAB bindings v1.14.0). Pump flow was obtained from the heart–lung machine either directly via its serial interface (RS-232) or as an export from the patient data management system (MetaVision, iMDsoft), depending on the operating theatre. As far as surgery allowed, manipulation of the patient, ultrasound and irrigation of the surgical field were suspended during indicator transit.

#### Filtering and channel selection

Each bolus recording was filtered with an adaptive procedure in in-house R code before fitting. It was designed to preserve the timing and height of the bolus: conventional filters with fixed cut-offs either delay the peak (group delay of recursive filters, which forward–backward filtering does not remove at steep bolus edges) or flatten it and shift its onset (uniform smoothing), and either would bias TTP, rise time and the amplitude ratio. Motion spikes of up to 0.5 s were detected against a rolling median (beyond 6 median absolute deviations when periodic, 10 otherwise) and repaired by interpolation. Physiological oscillations were removed per bolus rather than with fixed cut-offs. For each bolus, a master spectrum was formed as the median Welch power spectrum of all channels (window of 256 samples, 50% overlap), and spectral peaks were sought in the cardiac (0.5–2.0 Hz), respiratory (0.19–0.50 Hz) and two vasomotor bands (up to 0.13 and 0.13–0.19 Hz), with the lower vasomotor bound being kept above the time scale of the bolus itself. A band was filtered only where the power at its peak exceeded the spectral background of the band ($z \geq 3$) above the bolus time scale, with a zero-phase notch, applied as a Gaussian mask in the Fourier domain (width $f_0/Q$), placed within a window around the peak (cardiac ± 0.16 Hz with harmonics, Q 25; respiratory ± 0.11 Hz, Q 10; vasomotor Q 8). Cardiac notches were applied in 55 of 94 boluses (median 0.96 Hz), in 21 of 22 off CPB and 34 of 72 on CPB; respiratory notches were applied in 45 (median 0.24 Hz) and vasomotor notches in 5 and 11. On CPB, peaks in the cardiac and respiratory bands were also found during cardioplegic arrest (16 boluses in P3–P8), where they presumably arise from the roller pumps rather than from the heart or ventilation; the notch removes them irrespective of their source. Each channel was then smoothed with a weighted smoothing spline whose stiffness followed the bolus kinetics, being stiffer for slower passages (smoothing parameter 0.40–0.95 for provisional rise times of 2–35 s, estimated from the pre-smoothed trace with onset at 5% of its maximum), and with additional weight on a window around the first pass, so that slow intraoperative curves are smoothed strongly while the steep first-pass flank and its maximum keep their position and height, and resampled to 4 Hz from a recording rate of 5.1 Hz (6.1 Hz in the first two patients). A channel entered the fit if it showed a positive bolus deflection and a bolus signal-to-noise ratio of at least 100, defined as the maximum of the post-injection signal relative to the robust standard deviation of its sample-to-sample differences.

#### Channel model and channel yield

Each channel was analysed from 15 s before to 120 s after injection. Its pre-injection mean was subtracted as offset. For each bolus, an analysis window defined which samples entered the offset and the fit: it excluded all data before −15 s and, by default, all data after 60 s; where late recirculation or artefacts would otherwise enter the fit, the upper bound was set earlier based on visual inspection of the bolus (43 of 94 boluses, 15–59 s) and, in two boluses, later (65 and 80 s). The first pass was fitted with a Janoschek wash-in (Janoschek 1957) and a delayed exponential washout,

$$C(t) = A\left[1 - \exp\left(-g(t-\delta)^{s}\right)\right]\exp\left(-k\,\tau(t)\right), \tau(t) = \frac{1}{\beta}\ln\left(1 + e^{\beta(t-\delta-\lambda)}\right)$$

for $t > \delta$ (zero before), with amplitude $A$, delay $\delta$, growth rate $g$ and shape $s$ of the wash-in, washout rate $k$ after the lag $\lambda$, and a smooth onset of washout ($\beta = 1.5$). The fit window ends halfway between the steepest descent after the peak and the following inflection of the measured curve, i.e. before the onset of recirculation; parameters were estimated by Levenberg–Marquardt least squares, with starting values determined by a grid search instead of being set arbitrarily and a bounded quasi-Newton method (L-BFGS-B) as a fallback, and $R^2$ computed over the fit window. From the fitted curve on a 0.1-s grid, TTP is defined as the time of its maximum, the onset T0 as the first time it reaches 1% of that maximum, and the rise time as TTP − T0. Channel yield is the number of channels of a hemisphere passing both acceptance criteria ($R^2 \geq 0.90$, rise time < 59 s) divided by the number of montaged channels of that hemisphere at 40 and 54 mm. The montage was side-symmetric for every bolus, so both hemispheres share the denominator and their difference needs no correction for the number of available channels. Channel yield was analysed with the same mixed model as ΔTTP.

#### Reference configuration

The baseline before CPB (P1) is the only configuration with native circulation and among the least variable (SD of ΔTTP 3.5 s). The baseline on CPB (P2) is unsuitable as a reference, because its asymmetry is expected to vary between patients: the bolus enters through the right subclavian cannula and reaches the left-sided vessels only with the residual ejection of the left ventricle, which differs from patient to patient (Table S2). Accordingly, P2 is the most variable configuration in the sequence (SD 9.2 s), and as a reference level, it would carry that variance into every contrast.

#### Hemisphere-specific values

Where hemisphere-specific values are shown or ranked rather than contrasted (Fig. 3A and B), they were adjusted within the same framework: for each hemisphere and quantity, a model with configuration as fixed effect and a random intercept per patient was fitted, and the patient's offset, together with the model level of P1, was removed from each value. This removes the native side asymmetry a patient carries before any perfusion strategy is applied; raw values are reported alongside.

#### Multiplicity

Within each family of hypotheses, *p*-values were adjusted by the Holm method (Holm 1979). Eleven families comprising 89 tests are reported: the nine configuration contrasts of ΔTTP; the nine of the interhemispheric $rSO_2$ difference and the nine of the same quantity against its own reference configuration; the seven contrasts of absolute $rSO_2$ levels; the three post hoc steal contrasts on the raw and the adjusted scale; the three within-hemisphere contrasts behind them; the nine contrasts of channel yield and the nine of the same analysis without the extreme bolus; the ten configurations of the rank scan; and the two sets of nine contrasts comparing ΔTTP with $rSO_2$. No correction was applied between families, as the families are not of equal standing: the ΔTTP contrast family carries the primary analysis the study was designed for, with expectations specified for each configuration (Table S2), whereas the other families are only supporting or exploratory — and the ΔTTP finding does not rest on a single test: six of its nine contrasts survive Holm correction, five of them in the same direction.

#### Oximetry ceiling

The clinical oximeter displays $rSO_2$ up to 95%. Readings at that ceiling were retained; they bias associations with $rSO_2$ towards zero, i.e. towards a null effect, and their proportion is reported with each analysis.

#### Software

Preprocessing and model fitting ran in R 4.5.1, the statistical analysis and the figure creation were performed in R 4.5.3 (R Core Team 2026). Preprocessing used data.table 1.17.8 (Barrett 2025), zoo 1.8-14 for rolling medians (Zeileis 2005) and gsignal 0.3-7 for Welch spectra and notch filters (van Boxtel 2024); the channel model was fitted with minpack.lm 1.2-4 (Levenberg–Marquardt; Elzhov 2023) and pracma 2.4.4 (Borchers 2023), with the bounded fallback in the stats package. Statistics used lme4 2.0-6 (Bates 2015), lmerTest 3.2-1 (Kuznetsova 2017) and emmeans 2.0.4 (Lenth 2026), figures were created using ggplot2 4.0.3 (Wickham 2016).

### Supplementary results

#### Feasibility and channel attrition

The two boluses without a bilateral value came from one patient. In the reversal configuration (P7) the left-hemispheric signal collapsed (1.5% of left-sided channels retained against 55.9% on the right); in the preceding configuration (P6) both hemispheres fell below the acceptance criteria (0% left, 8.8% right). Across all boluses 22,432 channels were montaged and 15,948 (71.1%) met both acceptance criteria; at the 40- and 54-mm separations on which the analysis rests, this was 8,826 of 12,784 (69.0%). Most of the attrition occurred during channel selection, before the fit: of the 16,461 channels that entered the fit, 96.9% passed. This was asymmetrical between the hemispheres:

61.5% of the montaged left-hemispheric channels against 80.5% of the right-hemispheric channels entered the fit. Both causes of this asymmetry are systematic: interference with the clinical oximetry sensors and the left-dominant signal loss under SACP, which is a finding rather than a quality defect (see Channel yield across perfusion configurations).

#### Channel yield across perfusion configurations

The left hemisphere keeps fewer channels in every configuration (Figure S1); differences are given as right minus left, such that a positive value marks the left hemisphere as disadvantaged. Taken per bolus, the median side difference of a configuration lies between +7.4 points (rewarming) and +38.2 points (trilateral SACP, three boluses); these are medians of per-bolus differences, not the pooled proportions of Table S3, which put the largest difference at +32.4 points in that same configuration. The left hemisphere already keeps fewer channels before bypass (+13.2 points), an offset attributed to the clinical oximetry (see below). Against that baseline, no configuration is established: the only nominally significant contrast is +12.2 points under bilateral selective perfusion (+1.7 to +22.6, $p$ = 0.023, Holm 0.207), the largest, under trilateral perfusion (+15.3 points, three boluses), is a trend ($p$ = 0.075), and no contrast survives the correction. This is a limit of what these data can establish rather than evidence of absence: the nine intervals span −16.7 to +32.2 points, so configuration effects as large as the pre-bypass deficit itself remain compatible with them; furthermore, with three to fourteen boluses per configuration, the design lacks sufficient power to separate such effects from the scatter. What the analysis does establish is the asymmetry inherent in the recording at the start of the operation, which is present before bypass and which explains variance between patients rather than between configurations. One bolus is extreme and has been retained: under bilateral selective perfusion, one patient kept 7.4% of the left-sided against 92.6% of the right-sided channels, a raw difference of +85.3 points. Omitting this value shifts the single nominal finding rather than removing it — bilateral selective perfusion falls to +6.9 points ($p$ = 0.130) while trilateral perfusion reaches $p$ = 0.042, because the shared residual standard deviation drops from 12.7 to 10.7 points and the sparsest cell gains most from that; after Holm correction, neither version establishes any contrast.

Across configurations, the yield asymmetry follows the TTP asymmetry: on the scale of the two models, the configuration estimates correlate at Spearman $\rho$ = +0.70 (exact permutation $p$ = 0.031 over all 3,628,800 assignments of the ten configurations), the positive sign meaning that where the left hemisphere peaks later, it also keeps fewer channels. On the raw medians the same correspondence is weaker and not established ($\rho$ = +0.43, exact $p$ = 0.22). Both $p$-values are exact, obtained by enumerating the permutation distribution rather than sampling from it. The one configuration that deviates from this pattern is the reversal — unilateral selective perfusion with the left subclavian and the left common carotid artery on body perfusion — where the left hemisphere peaks 5.7 s *earlier* and still keeps 18.4 points fewer channels; both values are read on the scale of the figure (the modelled level of that configuration), not the contrast against the pre-bypass baseline reported in the main text (−6.99 s) nor the pooled channel proportions of Table S3 (+23.3 points). Excluding this configuration increases the coefficient to $\rho$ = +0.87 ($p$ = 0.005). This value is the extreme of a leave-one-out series and not a second result: omitting each configuration in turn yields $\rho$ values between +0.58 and +0.87, with exact $p$-values between 0.005 and 0.108. The coefficient fails to reach nominal significance whenever unilateral, bilateral or trilateral selective perfusion, the body-perfusion configuration P6, or the completed arch is left out — five of the ten omissions. With ten points the correspondence is therefore an ordering statement about the configurations and not an estimate of its strength. Read together with the TTP asymmetry in Figure 2, the yields and the underlying channel attrition follow the same side and the same ordering as the TTP effects, consistent with the proposed steal mechanisms, while the yield contrasts themselves are too noisy to carry that statement on their own.

The offset before bypass most likely reflects the clinical INVOS oximetry, which could not be omitted without compromising standard care. Its sensor cables were routed to the right, so that the sensors sat in an asymmetric geometry relative to the CE-NIRS optodes, with the INVOS light source lateral to the array on the left and medial to it on the right; higher left-sided dark noise and stray light from the operating lamps may have contributed. Because these sources were constant throughout the surgery, they cannot account for the configuration-dependent part of the yield difference, which widened under selective cerebral perfusion and returned towards baseline after arch completion, in parallel with ΔTTP.

#### Informative missingness

Channels drop out when no interpretable bolus passage reaches them, so the missingness carries information. In the configurations with a right-sided lead (P2 to P6) it preferentially removes late left-hemispheric passages, and the corresponding ΔTTP estimates are, if anything, conservative. In P7, where the left hemisphere peaked first but still lost more channels, the same selection could exaggerate the magnitude of the reversal.

#### Cerebral oximetry: configuration contrasts

The interhemispheric $rSO_2$ difference (right − left; positive = lower on the left) was analysed with the model used for ΔTTP. Against the pre-bypass baseline (raw median +3.95 percentage points) the nine contrasts were +1.9 (P2), +5.4 (P3), +2.4 (P4), +0.6 (P5), +0.1 (P6), −3.0 (P7), −0.7 (P8), −2.9 (P9) and −1.2 percentage points (P10). Only P3 reached nominal significance (95% CI +1.3 to +9.6, $p = 0.010$), and none survived Holm correction (P3 adjusted $p = 0.094$; all others 1.000). The direction of the P7 contrast, a relatively higher left-sided saturation, matches the reversal of ΔTTP, but its interval includes zero (−7.0 to +1.1).

#### Coupling of $rSO_2$ and TTP: sensitivity

The association reported in the main text (P4 to P8, configuration as fixed effect, random intercepts for patient and bolus) was refitted with random slopes for TTP per patient and per hemisphere. The left-sided slope remained between −0.236 and −0.291 percentage points per second and excluded zero in all three versions (patient slope −0.236, −0.429 to −0.044; slope per hemisphere −0.291, −0.500 to −0.082); the right-sided slope stayed between −0.209 and −0.215, with intervals reaching just above zero, and the two hemispheres did not differ in any version (interaction $p$ 0.37 to 0.71). These fits were singular, with random-slope variances at or close to the boundary, so with nine patients and 35 boluses, patient-specific slopes are not identified, and the intercept-only model is reported as primary. Without the configuration term, the slopes are flatter (left −0.118 to −0.188) and their intervals include zero: the coupling is a within-configuration relation and is not carried by level differences between configurations. Across all ten configurations, the left-sided slope is −0.232 (−0.350 to −0.114) and the right-sided slope is −0.140 (−0.275 to −0.005).

### Supplementary discussion

#### Feasibility in context

ICG bolus tracking with CE-NIRS has been applied during selective cerebral perfusion in piglets (Meybohm 2008) and in chronic occlusive cerebrovascular disease, where the side ratios of its kinetic parameters correlated with positron emission tomography (Saito 2018). To our knowledge, the present study represents the first application of this technique to the intraoperative monitoring of SACP in adult aortic arch repair, including the periods of hypothermia and circulatory arrest that make intraoperative neuromonitoring particularly demanding.

#### Rewarming

The left-sided delay expected for rewarming (P9; Table S2) was not confirmed (+1.0 s, −3.1 to +5.1). The expectation rested on the same mechanism as in P2, a bolus that reaches the left-sided vessels with the residual ejection of the left ventricle. Depending on the timing of the bolus during rewarming, left-ventricular ejection had probably not yet resumed to a relevant extent, so that the arch was supplied by the circuit alone and the bolus reached both sides alike.

#### Individual strength of the steals

Within a single configuration, TTP asymmetry varied widely between patients: from −1.9 to +24.8 s under bilateral SACP (P4), from +1.8 to +15.4 s with the left subclavian artery on body perfusion (P6), and from −8.8 to +1.9 s in the reversal configuration (P7). This is consistent with the configurations creating the condition for a steal, while its strength in a given patient depends on further factors, including collateral capacity, the geometry of the cannulation and, given the confounding described in the main text, the cannulation strategy itself. The separate SACP roller pump does not exclude the Y-steal: it is not run fully occlusively, so that it does not impose its flow on the SACP limb, and where the body-perfusion limb beyond the Y-connector offers the lower resistance, the pump keeps turning while delivering less to the SACP limb. Pump flow is not measured but calculated from pump speed, and only that of the arterial pump was documented, so the flow actually reaching the SACP limb could not be quantified.

#### Can longer paths from the T-connector explain the delay?

A longer line to one hemisphere delays the arrival of the bolus there, independently of any maldistribution of flow. Whether this alone can account for the observed asymmetry can be tested at the T-connector under bilateral SACP (P4). In this configuration, the geometry of both lines is known, and the documented arterial pump flow equals the SACP flow (median 1.67 L/min, 0.92–2.10, 12 boluses). For the Y-connector, the same test is not possible, because the flow in the selective and body-perfusion limbs beyond it was not measured. Beyond the T-connector, the line to the left common carotid artery (LCCA) is 71 cm long, and that to the right subclavian artery (RSA) is 10 cm. Assuming an inner diameter of 4.6 mm for both, their priming volumes are 11.8 and 1.7 mL, respectively. The common segment from the heart–lung machine to the T-connector is passed by the bolus on both sides and therefore cancels out. The transit time of a line is its priming volume divided by its flow. With the flow divided equally between the two lines, the longer line delays the left-sided bolus by 0.7 s (0.6–1.3), compared with an observed median of 6.9 s. Accounting for tubing resistance (Hagen–Poiseuille law, viscosity 3 mPa·s, the upper end of the expected range, left line including a 2-mm stopcock of 3.5 cm length) and an equal downstream resistance of the cannula and vascular bed, estimated as a perfusion pressure of 60 mmHg (Maier 2016) divided by half the SACP flow, changes the division little: the left line then carries 35–42% of the flow, and the delay is about 1 s (0.9–1.7). Explaining the observed delay by transit time alone would require the left line to carry only about 8% of the SACP flow, that is, a total resistance on the left more than ten times that on the right. Because the tubing contributes little to this difference, such a disparity can only arise further along the path. The cannulas are unlikely to account for it: the RSA line ends in an 8-mm graft, and a 15-Fr LCCA cannula (outer diameter 5 mm) with a plausible lumen of 3.5–4.5 mm raises the estimated delay only to about 1.1–1.2 s. The disparity therefore most likely lies in the vascular bed. Because flow in the individual lines was not measured, this remains to be confirmed.

#### A single trajectory through both proposed steals

Figure 3D shows the boluses of patient A, who sustained a left-hemispheric infarction, on the raw scale. Before bypass, both hemispheres behaved alike (ΔTTP +1.8 s, left-to-right amplitude ratio 0.97). Under bilateral SACP, the left hemisphere peaked 24.8 s later than the right, and its relative amplitude fell to 0.65. With the left subclavian artery on body perfusion (P6)—the configuration in which both steals may act on the left hemisphere—the ratio fell further to 0.28, the lowest value in this patient, despite a higher total pump flow (6.0 against 2.1 L/min); ΔTTP was +15.4 s, although only 9 left-hemispheric channels contributed. In the reversal configuration (P7), the sign inverted: the left hemisphere peaked 8.8 s ahead, and channel yield now dropped on the right (21% against 44% on the left), consistent with the Y-steal acting on the right hemisphere. During rewarming, the left hemisphere showed the higher amplitude (ratio 1.76), which may indicate reactive hyperaemia after the preceding deficit (Torre 2026).

#### Steal-specific saturation contrasts

The configurations in which we propose the steals to act were not associated with a clear saturation change in the hemisphere at risk. Each contrast compares the exposed configurations with the remaining ones from window P4 to P8 in the same hemisphere, rather than with the pre-bypass baseline: for the T-steal, the left hemisphere in P4 to P6 against P7 and P8 (−0.11 percentage points, 95% CI −4.26 to +4.04); for the Y-steal, the right hemisphere in P6 and P7 against P4, P5 and P8 (+0.84 percentage points, −4.61 to +6.28). Nor was the change specific to the hemisphere at risk: in a joint model of both hemispheres, the exposure × hemisphere interaction (right minus left) pointed in the predicted direction for both steals but was not significant (T-steal +2.54

percentage points, 95% CI −1.29 to +6.36, $p = 0.202$; Y-steal −1.52 percentage points, −5.78 to +2.74, $p = 0.488$). The device ceiling of the oximeter at 95% further limited the range of these contrasts. Together with the small within-configuration coupling of $rSO_2$ and TTP, this indicates that the two quantities capture different aspects of hemispheric supply: the kinetics of indicator delivery on the one hand, and a lagging, ceiling-limited balance of oxygen supply and consumption on the other.

#### Lesion site, recorded field and vascular anatomy

How closely a lesion can correspond to the measurement differs between the three patients (Table S5). The montage covers the middle cerebral artery territory, although only partially. In patient F, both the right-hemispheric infarction and the left-hemispheric oedema lie in that territory; thus, this represents the trajectory the method could, in principle, have tracked. A dissection membrane already extended into the left common carotid, subclavian and vertebral arteries before surgery — the vessels on which both steals act — so that the vascular abnormality here is a substrate that favours a steal rather than a consequence of the repair. In patient A, the infarcts lie in the borderzone between the middle and the posterior cerebral artery territories; with a fetal-type left posterior cerebral artery, that borderzone is supplied by the same internal carotid artery, so the lesion is downstream of the compromised path but at or beyond the edge of the recorded field. In patient G, the infarction lies in the anterior cerebral artery territory, which the montage does not cover; with a narrow right A1 segment and absent posterior communicating arteries, this territory depends on cross-flow through the anterior communicating artery rather than on the carotid we were measuring. Additionally, a suspected high-grade stenosis at the origin of the right common carotid artery, attributed radiologically to the anastomosis of the graft onto the brachiocephalic trunk, makes a mechanical cause a competing explanation. The correspondence between reversal and injury is thus anatomically plausible in one patient, partial in the second and absent in the third.

#### Laterality

The laterality of the injuries (one left-hemispheric, one right-hemispheric, one bilateral) argues against interpreting the reversal as a side-specific mechanism in which the later-peaking hemisphere is the one at risk. We instead propose that the magnitude of the reversal indexes how strongly the Y-steal acts in an individual patient, and that its consequences can affect either hemisphere depending on the configuration: on the right in P7, when the right hemisphere is supplied through the SACP limb alone, and on the left in P6, when the Y-steal adds to the T-steal at the left carotid cannula. Because the patients with cerebral injury were also those managed trilaterally — a strategy chosen because of the posterior circulation or variants of the circle of Willis — the magnitude of the reversal may also reflect limited collateral compensation, which would allow a steal to take full effect on the dependent hemisphere.

Three observations are consistent with this interpretation, to differing degrees. Patient A, with the most pronounced reversal, had his lowest left-hemispheric amplitude in P6 (ratio 0.28) and subsequently sustained a left-hemispheric infarction. Patient F, with bilateral injury and the second most pronounced reversal, showed both components: in P6, a marked left-hemispheric delay with the amplitude ratio falling to about one-fifth of its already reduced baseline value, consistent with the radiological attribution of the left-hemispheric oedema to hypoxia; and in P7, a pronounced right-sided delay and a drop in right-hemispheric channel yield (Figure S2A). The amplitude loss in P6 was confined to these two patients: among the five patients with a bilateral value in this configuration, the two with left-hemispheric injury had the lowest left-to-right amplitude ratios (0.08 and 0.28), whereas the other three, none of whom sustained postoperative cerebral injury, retained left-sided amplitude (0.64, 1.04 and 2.14); with two against three patients, the exact rank-sum test cannot fall below $p = 0.20$, so this observation is hypothesis-generating. Patient G, with right-hemispheric infarction, likewise showed a marked right-sided delay in P7 (−7.5 s), with a raw amplitude ratio of 1.08, but without the left-sided amplitude loss of the other two patients; across his remaining configurations, the ratio ranged from 0.37 to 0.99 (Figure S2B).

### Supplementary tables

**Table S1. Full patient and operative characteristics (*n* = 10).** Data are *n* (%) or mean ± SD. The cerebral perfusion strategy is counted per patient from the surgical protocol. AVR: aortic valve replacement; COPD: chronic obstructive pulmonary disease; CPB: cardiopulmonary bypass; FET: frozen elephant trunk; LVEF: left ventricular ejection fraction; TIA: transient ischaemic attack; VSARR: valve-sparing aortic root replacement.

| Characteristic | Value |
|---|---|
| **Demographics** | |
| Female sex | 3 (30%) |
| Age at surgery, years | 66.6 ± 10.7 |
| Height, cm | 174.9 ± 6.7 |
| Weight, kg | 86.3 ± 26.2 |
| Body mass index, kg/m² | 27.9 ± 7.0 |
| Body surface area, m² | 2.0 ± 0.3 |
| **Cardiovascular risk factors** | |
| Hypertension | 10 (100%) |
| Diabetes mellitus | 2 (20%) |
| Obesity | 1 (10%) |
| Smoking history | |
| Never smoker | 7 (70%) |
| Active smoker | 2 (20%) |
| Former smoker | 1 (10%) |
| **Cardiac status** | |
| Coronary artery disease | 1 (10%) |
| History of myocardial infarction | 0 (0%) |
| LVEF <50% | 2 (20%) |
| Preoperative atrial fibrillation | 2 (20%) |
| **Peripheral and cerebrovascular disease** | |
| Peripheral arterial disease | 1 (10%) |
| History of stroke | |
| No previous stroke | 9 (90%) |
| Previous stroke with residual deficits | 0 (0%) |
| Previous stroke without residual deficits | 1 (10%) |
| History of TIA | 1 (10%) |
| Supra-aortic vessel stenosis >65% | 4 (40%) |
| Supra-aortic vessel dissection | 3 (30%) |
| **Pulmonary and connective tissue disease** | |
| COPD/asthma | 2 (20%) |
| Connective tissue disorder | 0 (0%) |
| **Aortic pathology and surgical history** | |
| Diagnosis | |
| Native arch aneurysm | 4 (40%) |
| Post-dissection aneurysm | 1 (10%) |
| Postoperative aneurysm progression | 5 (50%) |
| Previous aortic surgery | 6 (60%) |
| Previous type A dissection | 1 (10%) |
| Previous root replacement | 1 (10%) |
| **Operation** | |
| Isolated FET | 3 (30%) |
| Additional aortic procedure | |
| None | 5 (50%) |
| AVR | 0 (0%) |
| Conduit | 2 (20%) |
| VSARR | 1 (10%) |
| Ascending replacement | 2 (20%) |
| Additional cardiac procedure | 5 (50%) |
| Number of redo cardiac procedures | |
| 0 | 3 (30%) |
| 1 | 6 (60%) |
| 2 | 1 (10%) |
| Operative urgency | |
| Elective | 8 (80%) |
| Urgent | 2 (20%) |
| Emergency | 0 (0%) |

| Characteristic | Value |
|---|---|
| Cerebral perfusion strategy | |
| Unilateral | 0 (0%) |
| Bilateral | 6 (60%) |
| Trilateral | 4 (40%) |
| EuroSCORE II, % | 5.7 ± 4.8 |
| Operation time, min | 404.2 ± 80.7 |
| CPB time, min | 237.5 ± 47.7 |
| Cross-clamp time, min | 118.9 ± 30.2 |
| Lowest body temperature, °C | 24.4 ± 2.1 |

**Table S2.** Perfusion configurations traversed during frozen elephant trunk repair and the TTP asymmetry expected for each of them from its intended function. Side difference is defined as the left minus the right hemispheric value of the bolus TTP, so a positive value means the bolus reaches the right hemisphere earlier. Phases are numbered in surgical order.

| Phase | Perfusion configuration | ICG injection | Expected side difference in TTP | Expectation | Rationale |
|---|---|---|---|---|---|
| P1 | Baseline, before cardiopulmonary bypass | central venous | none | no side differences | Native circulation, symmetric supply of both carotid arteries |
| P2 | Baseline, on cardiopulmonary bypass | bypass circuit | left later | slightly delayed, more dispersed left-sided bolus passage (dependent on residual LV ejection; altered dispersion/dilution, preserved flow) | Cannula in the right subclavian artery, retrograde into the brachiocephalic trunk; the left hemisphere is reached over the arch from the residual cardiac ejection and is therefore further diluted and dispersed |
| P3 | Unilateral SACP (right subclavian artery) | bypass circuit | left later | delayed, more dispersed left-sided bolus passage (driven by collateralisation) | Injection into the right subclavian artery; the left hemisphere is supplied through the circle of Willis only |
| P4 | Bilateral SACP (right subclavian + left common carotid) | bypass circuit | none | no side differences, symmetry restored by SACP | The left common carotid artery is cannulated, so the anterior and middle cerebral artery territories of both hemispheres are perfused directly; the posterior territories depend on the circle of Willis |
| P5 | Trilateral SACP (right subclavian + left common carotid + left subclavian or vertebral) | bypass circuit | none | no side differences, symmetry restored by SACP | The left subclavian artery is cannulated in addition, so all arch vessels are perfused directly |
| P6 | Bilateral SACP, left subclavian on body perfusion | bypass circuit | none | no side differences, symmetry restored by SACP | Left subclavian artery incorporated into body perfusion, right subclavian and left common carotid on the selective line |
| P7 | Unilateral SACP, left subclavian and left common carotid on body perfusion | bypass circuit | none | no side differences, symmetry restored by SACP | Left subclavian and left common carotid artery on body perfusion, which runs at higher flow; right subclavian alone on the selective line |
| P8 | Arch open, all arch vessels on body perfusion | bypass circuit | none | no side differences | All arch vessels incorporated into body perfusion |
| P9 | Rewarming | bypass circuit | left later | slightly delayed, more dispersed left-sided bolus passage (dependent on reinitiated LV ejection; altered dispersion/dilution, preserved flow) (as in P2) | Body perfusion at full flow, and the heart ejects again, so the left hemisphere is once more reached over the arch from the ejected blood |
| P10 | End of surgery | central venous / peripheral venous | none | no side differences | Restored circulation, injection venous again |

**Table S3.** Recorded boluses, patients and channel yield per perfusion configuration. The configurations follow the course of the operation; the specific configurations to which a patient contributes are determined by the cannulation strategy and the individual surgical course, so the number of patients differs between configurations. A bolus counts as having a bilateral time to peak (TTP) when the acceptance criteria were met on both sides; only those boluses enter the side-difference models. The channel yield is the proportion of montaged channels at 40 and 54 mm that passed the acceptance criteria, pooled across the boluses of the configuration; the difference is calculated as right minus left, such that a positive value, as for TTP, marks the left hemisphere as disadvantaged. The denominator of the patient count is the number of patients for whom the configuration was applicable: trilateral cannulation (P5) was performed in four patients, of whom three contributed a bolus in that configuration, so its coverage reads three out of four; every other configuration was possible in all ten patients and therefore counts against ten. Single boluses scatter widely around these proportions (standard deviation 22.5 pp on the left and 21.9 pp on the right, and 20.4 pp within a configuration in the median), so the two side values should be read as the pooled proportion rather than as precise per-bolus values.

| Phase | Perfusion configuration | Patients | Boluses | With bilateral TTP | Channel yield, left % | Channel yield, right % | Difference (right − left), pp |
|---|---|---|---|---|---|---|---|
| P1 | Baseline, before cardiopulmonary bypass | 9 of 10 | 11 | 11 of 11 | 61.5 | 77.9 | +16.4 |
| P2 | Baseline, on cardiopulmonary bypass | 10 of 10 | 14 | 14 of 14 | 56.9 | 78.8 | +21.8 |
| P3 | Unilateral SACP (right subclavian artery) | 7 of 10 | 7 | 7 of 7 | 63.7 | 87.6 | +23.9 |
| P4 | Bilateral SACP (right subclavian + left common carotid) | 10 of 10 | 13 | 13 of 13 | 57.1 | 82.8 | +25.7 |
| P5 | Trilateral SACP (right subclavian + left common carotid + left subclavian or vertebral) | 3 of 4 | 3 | 3 of 3 | 52.0 | 84.3 | +32.4 |
| P6 | Bilateral SACP, left subclavian on body perfusion | 6 of 10 | 6 | 5 of 6 | 32.6 | 54.4 | +21.8 |
| P7 | Unilateral SACP, left subclavian and left common carotid on body perfusion | 7 of 10 | 7 | 6 of 7 | 42.6 | 66.0 | +23.3 |
| P8 | Arch open, all arch vessels on body perfusion | 10 of 10 | 11 | 11 of 11 | 67.9 | 77.7 | +9.8 |
| P9 | Rewarming | 6 of 10 | 11 | 11 of 11 | 72.5 | 85.0 | +12.6 |
| P10 | End of surgery | 10 of 10 | 11 | 11 of 11 | 66.6 | 82.8 | +16.2 |

**Table S4.** Interhemispheric difference in bolus time to peak (TTP) per perfusion configuration. Values are left minus right in seconds, so a positive value means the bolus reached the right hemisphere earlier. Contrasts and intervals come from a linear mixed model with the phase as a fixed effect and a random intercept per patient, fitted on all boluses with a bilateral value; the baseline before cardiopulmonary bypass (P1) served as the reference. *p*-values are adjusted by the Holm method across the nine contrasts. The expected side difference corresponds to the expectations in Table S2.

| Phase | Perfusion configuration | Boluses (patients) | Median difference in TTP, s (IQR) | Boluses with left earlier | Expected side difference | Contrast against P1, s (95% CI) | p, Holm adjusted | Interpretation |
|---|---|---|---|---|---|---|---|---|
| P1 | Baseline, before cardiopulmonary bypass | 11 (9) | +1.70 (1.80) | 2 of 11 | none | reference | - | reference phase of the model |
| P2 | Baseline, on cardiopulmonary bypass | 14 (10) | +3.10 (14.12) | 0 of 14 | left later | +7.83 (+4.10 to +11.56) | 0.001 | as expected, direction confirmed |
| P3 | Unilateral SACP (right subclavian artery) | 7 (7) | +7.90 (7.65) | 1 of 7 | left later | +5.95 (+1.42 to +10.48) | 0.043 | as expected, direction confirmed |
| P4 | Bilateral SACP (right subclavian + left common carotid) | 13 (10) | +9.00 (7.80) | 1 of 13 | none | +7.22 (+3.41 to +11.02) | 0.003 | departure from the expected symmetry, right earlier |
| P5 | Trilateral SACP (right subclavian + left common carotid + left subclavian or vertebral) | 3 (3) | +14.10 (6.05) | 0 of 3 | none | +8.69 (+2.54 to +14.84) | 0.031 | departure from the expected symmetry, right earlier; underpowered (n=3 patients) |
| P6 | Bilateral SACP, left subclavian on body perfusion | 5 (5) | +9.25 (2.85) | 0 of 5 | none | +7.86 (+2.75 to +12.97) | 0.021 | departure from the expected symmetry, right earlier |
| P7 | Unilateral SACP, left subclavian and left common carotid on body perfusion | 6 (6) | -6.20 (5.78) | 5 of 6 | none | -6.99 (-11.70 to -2.29) | 0.025 | departure from the expected symmetry, left earlier |
| P8 | Arch open, all arch vessels on body perfusion | 11 (10) | -0.10 (2.88) | 6 of 11 | none | -0.10 (-4.05 to +3.86) | 1.000 | no detectable difference |
| P9 | Rewarming | 11 (6) | +0.60 (5.90) | 5 of 11 | left later | +1.02 (-3.07 to +5.11) | 1.000 | expected delay not confirmed, interval includes zero |
| P10 | End of surgery | 11 (10) | +1.10 (2.17) | 4 of 11 | none | +0.07 (-3.90 to +4.04) | 1.000 | no detectable difference |

**Table S5. Patients with postoperative cerebral injury.** Clinical entries are taken from the radiology reports, the cannulation from the surgical protocol. A finding is defined as pre-existing if it was already evident on preoperative angiography, and as a consequence if the preoperative study showed no obstruction at that site; a mechanical complication is listed where it was established clinically. Patients are identified by the letters used throughout. The CE-NIRS montage covers the middle cerebral artery territory only partially, which is what the last column refers to.

| Patient | Cannulation | Vascular status before surgery | Cerebral lesion after surgery | New vascular finding after surgery | Substrate or consequence | Lesion within the CE-NIRS field |
|---|---|---|---|---|---|---|
| A | trilateral | thin mixed plaques of the left common carotid artery, no relevant stenosis; chronic occlusion of the right vertebral artery (distal V2), posterior circulation supplied by the left vertebral; complete circle of Willis; fetal-type left posterior cerebral artery | borderzone infarcts between the left middle and posterior cerebral artery territories (middle frontal gyrus, precuneus, cuneus) | no mechanical complication established | — | partly: the montage covers the middle cerebral artery territory, and the borderzone and occipital lesions lie at or beyond its edge |
| F | trilateral | dissection membrane extending into the LEFT common carotid artery (fading at the middle to distal third), the left subclavian and the proximal left vertebral artery; hypoplastic right vertebral artery, basilar supplied from the left; anterior communicating artery present | extensive right middle cerebral artery infarction including the basal ganglia, 12 mm midline shift; extensive left-hemispheric oedema in the middle cerebral artery territory, attributed to hypoxia | not assessed — the postoperative examination was an unenhanced CT | pre-existing substrate | yes: both the infarction and the oedema lie in the middle cerebral artery territory |
| G | trilateral | dissection membrane extending into the brachiocephalic trunk and the right subclavian artery with a dissecting aneurysm of the trunk, supra-aortic origins themselves unobstructed; no posterior communicating artery on either side; narrow right A1 segment; suspected stenosis of the intracranial right vertebral artery | infarction in the right ANTERIOR cerebral artery territory (parafalcine frontal to the central sulcus, later extending to the centrum semiovale); 17 days later, a large right cerebellar haemorrhage under anticoagulation | suspected high-grade ostial stenosis of the right common carotid artery at the anastomosis of the graft onto the brachiocephalic trunk, at the level of the carotid origin — a new finding compared with the preoperative study | consequence of the repair | no: the infarction lies in the anterior cerebral artery territory, outside the montage |

#### Supplementary figures

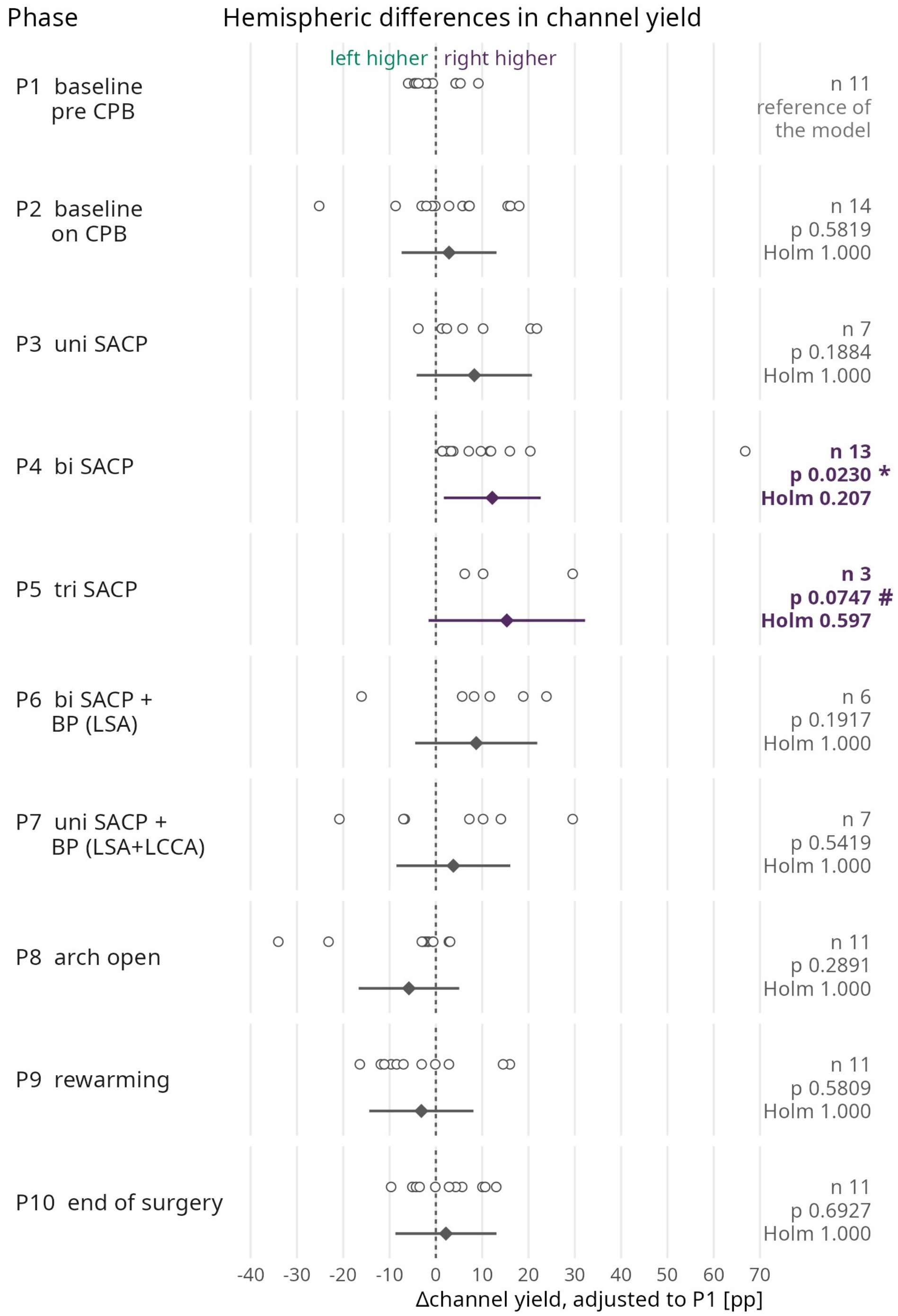


**Figure S1. Hemispheric difference in channel yield across the ten perfusion configurations.** Rows represent the configurations in surgical order. The yield of a bolus and hemisphere is the share of the montaged channels at 40 and 54 mm source–detector separation that satisfied both the goodness-of-fit and the rise-time criteria; because the montage was side-symmetric in every bolus, the two hemispheres share a denominator and their difference needs no correction for the number of available channels. Each open point is one bolus, drawn on the scale of a linear mixed model with configuration as a fixed and patient as a random effect and the pre-bypass baseline as the reference level — the same specification used in the main text for the TTP and for the interhemispheric $rSO_2$ difference — that is, with the patient offset and the reference level removed. The diamond below the points is the contrast of the configuration against the pre-bypass baseline with its 95% interval, *p*-values are two-sided and Holm-corrected over the nine contrasts. Zero on this axis is therefore each patient's own pre-bypass level and not the absence of an asymmetry: the persistent left-sided deficit sits in the reference level of the model (+14.7 percentage points) and in the patient offsets (−20 to +41 points), so the baseline row shows the residual scatter around that level rather than the deficit itself, which the Supplementary results above give as a raw median. As in the $rSO_2$ column of Figure 2, the difference is calculated as right minus left, such that a value to the right of zero marks the left hemisphere as the disadvantaged side, here the side that keeps fewer channels.

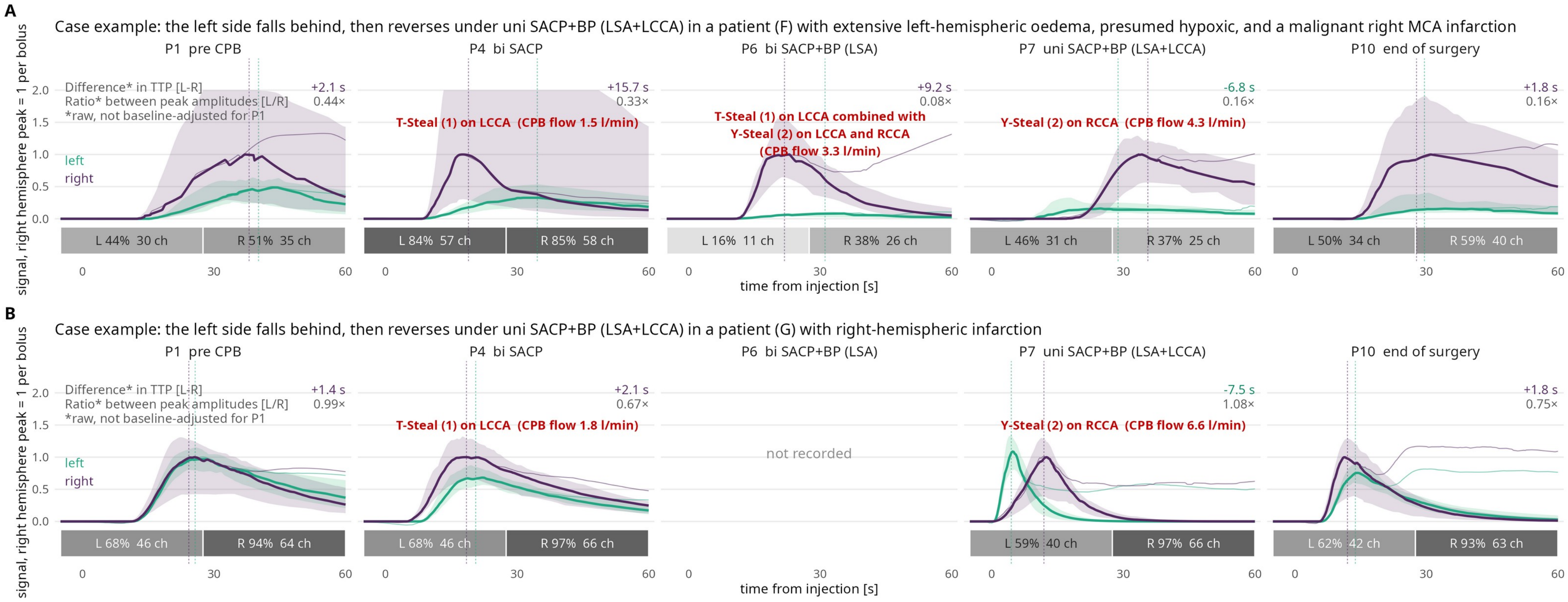


**Figure S2. Bolus series of the two remaining patients with postoperative cerebral injury.** Same construction as panel D of Figure 3. **(A)** patient F, with a malignant right middle cerebral artery infarction together with extensive left-hemispheric oedema of presumed hypoxic origin; **(B)** patient G, with a right-hemispheric infarction. Curves are the median of the fitted curves per hemisphere with interquartile band, scaled so that the right-hemispheric peak of each bolus is one; thin lines, measured signal; dashed lines, hemispheric TTP. The two numbers per facet are the measured TTP difference (left minus right) and the ratio of peak heights (left to right), both raw. The bar below each facet gives the proportion of montaged channels that passed the acceptance criteria, per side. The ordinate runs to 2 in both rows, clipping band tops in row A.
