## Supplementary material for "Near-infrared spectroscopy reveals asymmetric hemispheric blood supply during aortic arch surgery": STROBE Checklist

### STROBE checklist (cohort studies)

Locations refer to the section headings of the submitted manuscript. Checklist items abridged from the STROBE Statement for cohort studies (von Elm et al., 2007).

| Section | Item | Recommendation (abridged) | Reported in | Status |
|---|---|---|---|---|
| Title and abstract | 1a | Indicate the study design with a commonly used term in the title or the abstract | Abstract, Methods: “prospective, offline-analysed pilot study” | yes |
| | 1b | Informative and balanced summary of what was done and found | Abstract | yes |
| Background/rationale | 2 | Scientific background and rationale | Introduction | yes |
| Objectives | 3 | Specific objectives, including prespecified hypotheses | Introduction (last paragraph); expectations per configuration in Methods (Surgical technique and perfusion configurations) and Table S2 | yes |
| Study design | 4 | Key elements of study design early in the paper | Methods, Study design and patients | yes |
| Setting | 5 | Setting, locations, relevant dates (recruitment, exposure, follow-up, data collection) | Methods, Study design and patients (single centre, Freiburg, September 2022 to October 2024) | yes (outcome until discharge, Methods, Outcome) |
| Participants | 6a | Eligibility criteria, sources and methods of selection; methods of follow-up | Methods, Study design and patients (inclusion: aortic arch surgery with SACP; exclusion criteria); Outcome | yes (enrolled when the measurement team and equipment were available) |
| | 6b | Matched studies: matching criteria | — | not applicable |
| Variables | 7 | Define outcomes, exposures, predictors, potential confounders and effect modifiers; diagnostic criteria | Methods: perfusion configurations (exposure), ΔTTP and $rSO_2$ difference (endpoints), Outcome (cerebral injury); Results/Discussion: confounding by cannulation strategy | yes |
| Data sources/measurement | 8 | Sources of data and methods of assessment for each variable | Methods: Multichannel CE-NIRS and indicator protocol, Signal analysis, Outcome; Supplementary Methods | yes |
| Bias | 9 | Efforts to address potential sources of bias | Methods: specified expectations, acceptance criteria, patient random intercept; Limitations: imaging only in symptomatic patients, post hoc outcome comparison, reduced yield by oximetry | partly: no explicit statement, addressed in Limitations |
| Study size | 10 | How the study size was arrived at | Methods, Study design and patients (pilot, no formal sample size calculation) | yes |
| Quantitative variables | 11 | How quantitative variables were handled; groupings | Methods: Signal analysis (TTP, ΔTTP sign convention), Statistical analysis | yes |
| Statistical methods | 12a | All statistical methods, including control for confounding | Methods, Statistical analysis; Supplementary Methods | yes |
| | 12b | Methods to examine subgroups and interactions | Methods, Statistical analysis (TTP × hemisphere interaction; P7 comparison by outcome) | yes |
| | 12c | How missing data were addressed | Methods, Statistical analysis (patient without P1 bolus kept via reference level); Results, Feasibility (2 boluses without bilateral TTP; uneven coverage, Table S3) | yes |
| | 12d | Cohort: how loss to follow-up was addressed | Results, Feasibility (all ten patients completed the protocol) | yes |
| | 12e | Sensitivity analyses | Supplementary Methods and Supplementary Results | yes |
| Participants | 13a | Numbers at each stage (eligible, examined, confirmed eligible, included, completing follow-up, analysed) | Methods (10 enrolled); Results, Feasibility (10 completed, 94 boluses, 92 analysed) | partly: number screened/eligible not reported |
| | 13b | Reasons for non-participation at each stage | — | no (see 13a) |
| | 13c | Consider a flow diagram | — | no; optional |
| Descriptive data | 14a | Characteristics of participants; information on exposures | Table 1; Table S1; Methods (6 bilateral, 4 trilateral) | yes |

| Section | Item | Recommendation (abridged) | Reported in | Status |
|---|---|---|---|---|
| | | and potential confounders | | |
| | 14b | Number of participants with missing data for each variable | Table S3 (coverage per configuration); Results, Feasibility | yes |
| | 14c | Cohort: summarise follow-up time | Methods, Outcome (neurological symptoms until discharge) | yes |
| Outcome data | 15 | Cohort: numbers of outcome events or summary measures over time | Results, Postoperative cerebral injury (3 of 10); Table S5 | yes |
| Main results | 16a | Unadjusted and adjusted estimates with precision (95% CI); confounders adjusted for | Results: raw and P1-adjusted ΔTTP, contrasts with 95% CI; Fig. 2, Table S4 | yes |
| | 16b | Category boundaries when continuous variables were categorised | Methods/Results: ranking in P7, groups with and without injury | yes |
| | 16c | Absolute risk for a meaningful time period | — | not applicable |
| Other analyses | 17 | Other analyses (subgroups, interactions, sensitivity) | Results: Cerebral oximetry, Channel yield; Supplementary Results | yes |
| Key results | 18 | Summarise key results with reference to objectives | Discussion, Principal findings | yes |
| Limitations | 19 | Limitations, sources of bias or imprecision; direction and magnitude | Discussion, Limitations; Cerebral injury as a hypothesis; Results (ceiling of $rSO_2$ biases slopes towards zero) | yes |
| Interpretation | 20 | Cautious overall interpretation considering objectives, limitations, multiplicity, similar studies | Discussion, Flow competition, Comparison with cerebral oximetry, Cerebral injury as a hypothesis | yes |
| Generalisability | 21 | External validity | Discussion, Limitations (small single-centre cohort); Conclusions (prospective multicentre study) | partly: implicit |
| Funding | 22 | Source of funding and role of funders | Funding statement (funder had no role) | yes |
